# Epidemic prevention and control measures in China significantly curbed the epidemic of COVID-19 and influenza

**DOI:** 10.1101/2020.04.09.20058859

**Authors:** Xiang-Sha Kong, Feng Liu, Hai-Bo Wang, Rui-Feng Yang, Dong-Bo Chen, Xiao-Xiao Wang, Feng-Min Lu, Hui-Ying Rao, Hong-Song Chen

## Abstract

At the end of 2019, an outbreak of unknown pathogen pneumonia occurred in China, then it was named corona virus disease 2019 (COVID-19). With the rapid spread of COVID-19, a series of strict prevention and control measures were implemented to cut the spread of the epidemic. Influenza as a respiratory tract infection disease as COVID-19 might also be controlled. To assess the effects, we used the total passenger numbers sent in mainland China from 2018 to 2020 and the daily number of railway passenger (DNRP) flow in 2020 during Spring Festival travel rush to reflect the population movement and further to analyze newly and cumulative confirmed COVID-19 and influenza. We found that with implementing the series measures on COVID-19, not only COVID-19, but also influenza mitigated in China. The prevention and control measures for COVID-19 might be used in controlling respiratory tract diseases, and reducing the national health economic burden. When other countries issue measures on COVID-19 and influenza, they should consider adopting more aggressive epidemic prevention and control strategies.

---

At the end of December 2019, an outbreak of unknown cause pneumonia occurred in Wuhan, China^1^. The pathogen of the pneumonia had been identified as a novel coronavirus (2019-nCoV) on 7 January, 2020, which is now named severe acute respiratory syndrome coronavirus 2 (SARS-CoV-2) by the Coronavirus Study Group^2^, and the pneumonia is named corona virus disease 2019 (COVID-19) by World Health Organization (WHO) ^3^. On 19 January, the National Health Commission (NHC) listed COVID-19 caused by SARS-CoV-2 as Grade B infectious diseases, however, SARS-CoV-2 would be treated as a Grade A infectious disease, which requires the strictest prevention and control measures^4^.

In order to control the rapid spread of COVID-19, on 23 January, 2020, Wuhan was locked down and China raised the national public health response to the first state of emergency – the first level of 4 levels of severity in the Chinese Emergency System^5^. Then a series of severe prevention and control measures were implemented to curb the spread of the epidemic. Main measures of first-level response were displayed (Supplementary Table 1). Among them, to strictly limit population flow is a key measure to mitigate the spread of virus, the epidemic overlaps with the peak period of Spring Festival travel rush; therefore, reducing population movement might help to prevent the spread of COVID-19.

**Table 1.** Total passenger numbers sent by railway, highway, waterway and civil aviation during the Spring Festival travel rush period through 2018 to 2020

|  | 2020 | 2019 | 2018 | Contemporaneous<br>comparison<br>(2020 VS. 2019) | Contemporaneous<br>comparison<br>(2020 VS. 2018) |
| --- | --- | --- | --- | --- | --- |
| **Railway (100 million)** | 2.1 | 4.1 | 3.8 | 48.4% | 45.0% |
| **Highway (100 million)** | 12.1 | 24.6 | 24.8 | 50.8% | 51.2% |
| **Waterway (10 thousand)** | 1689 | 4077 | 4322 | 58.6% | 60.9% |
| **Civil aviation (10 thousand)** | 3839 | 7288 | 6541 | 47.3% | 41.3% |
| **Total (100 million)** | 14.8 | 29.8 | 29.7 | 50.5% | 50.3% |

Just like COVID-19,influenza is also a respiratory infectious disease, it could cause estimated 3-5 million cases with severe respiratory infection-related illness and 0.29-0.65 million deaths worldwide annual year^6,7^. And Influenza virus is easy to mutate and highly contagious, and could cause seasonal epidemic every year, mostly in winter and spring^8^. At the end and beginning of each year is influenza prevalence season.

This study mainly analyzes the influence of national first-level prevention and control measures on the epidemic of COVID-19 and influenza, and hope to provide some insights in curbing the COVID-19, influenza and other respiratory infectious diseases for other countries.

## Results

### Population flow dropping significantly since launching first-level response for the COVID-19

The Spring Festival travel rush period is a national transportation peak arranged by the Ministry of transport and the Civil Aviation Administration. Centering on the Spring Festival, it lasts 40 days, from the 15th of the 12th lunar month to the 25th of the first lunar month of the next year. It was from 10 January to 18 February in 2020 (25 January is the Spring Festival), from 21 January to 1 March in 2019, from 1 February to 12 March in 2018, respectively. In general, the Spring Festival travel rush period refers to intercity transportation in mainland China, including national railways, highway, waterways and civil aviation, and among them, national railways and highway are mainly transportation. We employ the daily number of railway passengers (DNRP) flow in mainland China to reflect the population movement and the speed and effectiveness of the first-level response to the epidemic.

During Spring Festival travel rush period in 2020, the total 1.48 billion passengers had been sent by railways, highway, waterway and civil aviation, decreased 50.5% and 50.3% compared to the same period in 2019 and 2018 respectively (Table 1). Among them, 210 million passengers sent by railway in 2020, which were 48.4% lower than the same period in 2019 and 45.0% lower than in 2018.

On 10 January, 2020, the DNRP in China had reached more than 10 million, at a high level from 10·49 to 12·44 million. According to previous data, the railway passenger flow showed a rapid downward trend two days before new year during 2018 to 2020 (Figure 1a). On 23 January, 2020, the DNRP flow was 9.85 million, higher than that of the same period in 2019 (6.0%) and 2018 (11.5%) respectively. At the same day, Wuhan was locked down and government launched a first-level response for the COVID-19. Therefore, on 24 January (the first day of lock down), the flow dropped to 5.15 million, lower than that of the same period in 2019 (6.0%) and 2018 (2.8%) respectively; and 2.47 million on 25 January, lower than that of the same period in 2019 (41.5%) and 2018 (36.6%) respectively. Furthermore, the daily railway flow dropped rapidly to 0.83 million on 13 February of 2020.

**Fig. 1.**
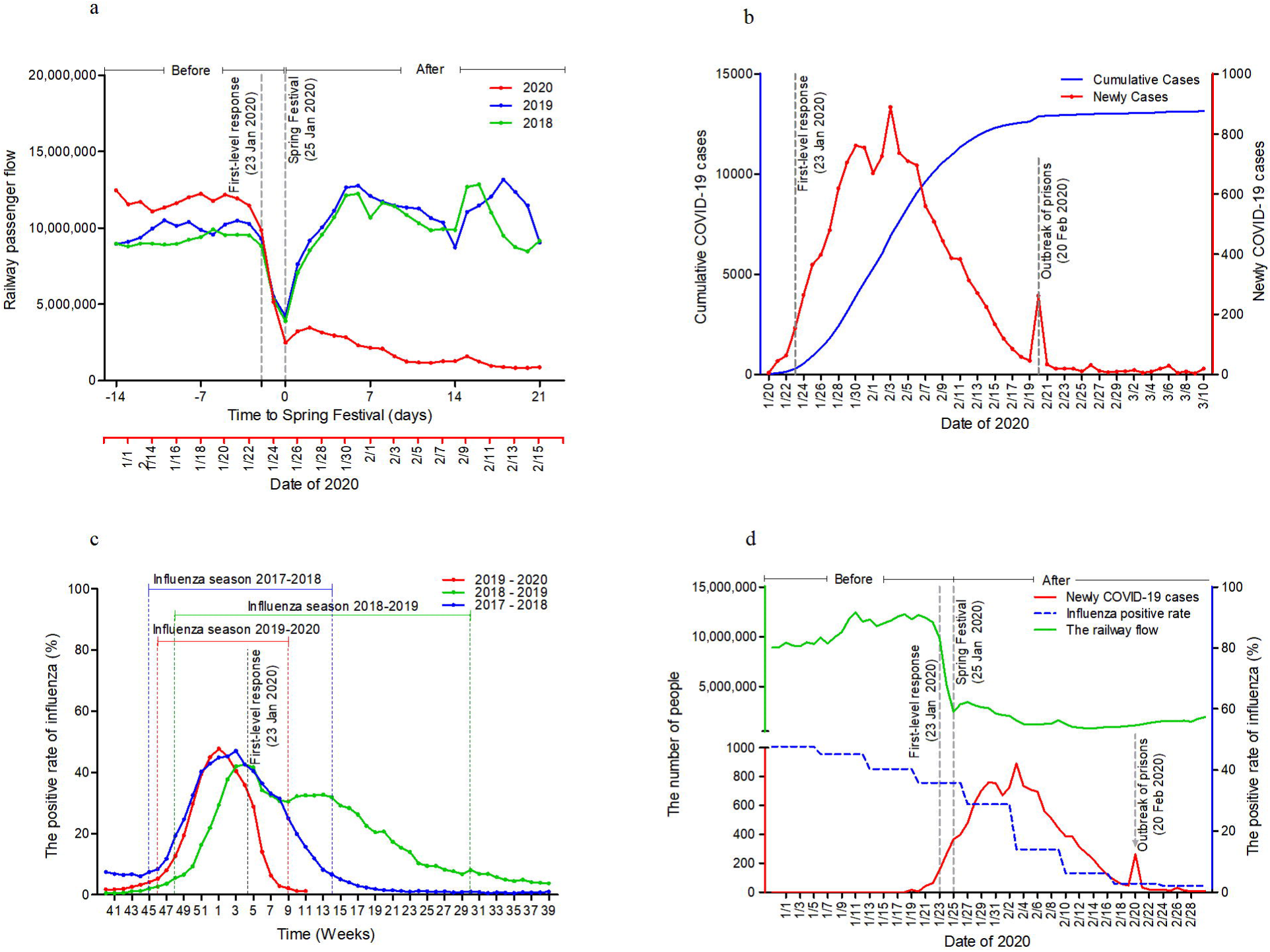
The effect of first-level response on the trends of railway passenger flow and the epidemic of COVID-19 and influenza. (a) The trends of railway passenger flow in Spring Festival travel rush period through 2018 to 2020. (b) The number of newly and cumulative COVID-19 cases tended to ease after 3 February. There was a little outbreak in the prisons of China’s Shandong Province and Zhejiang Province on 20 February. (c) All the data of influenza were observed using weekly data from the first week in October to the last week in September of next year. The trends of influenza epidemic seasons are very different in the latest three years. The space between two dashed lines of the same color represents the influenza season; red represents the influenza season of 2019-2020, green represents 2018-2019, and blue represents 2017-2018. (d) The influenza data derived from the weekly data reported by Chinese National Influenza Center (CNIC). In order to facilitate the observation of the daily dynamics of the epidemic prevention and control measures on population mobility and epidemic development, the daily positive rate of influenza was calculated by weekly data published from the influenza surveillance network laboratory of the mainland Chinese.

Based on data of 2018 and 2019, after the Spring Festival the DNRP flow quickly recovered to the high level. While in 2020, the daily flow in the first, second and third weeks after the Spring Festival were only 26.6%, 13.0% and 8.7% of the same period in 2019, respectively; and were only 28.3%, 13.3% and 10.0% in 2018.

### COVID-19 epidemic mitigated dramatically since launching first-level response for the COVID-19

After the first-level response prevention and control measures launched, the newly confirmed cases of COVID-19 still had an upward trend in other provinces and cities in China except Hubei province, then peaked on 3 February (Figure 1b). This temporary rise may be related with COVID-19 patients have traveled from Wuhan to other provinces and cities. Therefore, the outbreak has continued to spread geographically, with mounting numbers of cases and deaths. After 3 February, the number of newly confirmed cases in other provinces in China except Hubei province began to drop eventually. Until to 21 February, the number of newly confirmed cases was less than 50.

#### 2019-2020 Influenza epidemic season shortened in China

According to the epidemic data of influenza released by the Chinese National Influenza Center (CNIC), the 2019-2020 influenza season starts from the 46th week of 2019 to the 9th week of 2020, with the peak in the first week of 2020. The 2018-2019 influenza season starts from the 48th week of 2018 to the 30th week of 2019, with the peak in the third week of 2019. The 2017-2018 influenza season starts from the 45th week of 2017 to the 14th week of 2019, with the peak in the 4th week of 2018. The duration of 2019-2020 influenza season lasted only 15 weeks, which was significantly shorter than that of 2017-2018 (21 weeks) and 2018-2019 (34 weeks) (Figure 1c). In the 2019-2020 influenza season, the influenza-like illness (ILI) decreased to 59.2% of 2018-2019 and 64.0% of 2017-2018, respectively, and the influenza positive rate decreased to 82.8% of 2018-2019 and 78.1% 2017-2018 respectively.

One week before the first-level response on 23 January of 2020 (1/13-1/19), the positive rate of influenza was 40.4%. Then the positive rate of influenza in 1/20-1/26 reduced to 35.8%, and the following week (1/27-2/2) reduced to 28.7%. The positive rate in 2/24-3/1 rapidly reduced to 2.1%.

In 2019-2020, the time from the peak of the influenza positive rate to the Chinese first-level public health emergency response was 4 weeks. Accordingly, “the boundary point” was defined as 4 weeks added to the time when the influenza positive rate peaked in 2018-2019 and 2017-2018, which was the 8th week in 2018-2019 and the 7th week in 2017-2018, respectively. After “the boundary point”, the ILI in 2019-2020 decreased rapidly to 23.3% in 2018-2019 and 55.7% in 2017-2018. The influenza patients in 2019-2020 dropped to 11.0% and 31.6% compared with that in 2018-2019 and 2017-2018, respectively. The Influenza-A dropped to 13.1% and 26.9% compared with that 2018-2019 and 2017-2018, while Influenza-B decreased by 9.2% and 40.5%, respectively. (Figure 2 and Table 2)

**Table 2.** Comparison of the positive rate of influenza after the First Level Response with that before the response and the same period in 2019 and 2018

|  |  | Monitoring, No | Positive, No. | Influenza- A, No. | Influenza-B, No. |
| --- | --- | --- | --- | --- | --- |
| 2019-2020 | Before | 103677 | 32407 | 23771 | 8636 |
|  | After | 29351 (28.3%)* | 3157 (9.7%)* | 1763 (34.4%)* | 1394 (7.4%)* |
|  | Sum | 133028 | 35564 | 25534 | 10030 |
| 2018-2019 | Before | 98434 | 26929 | 26609 | 320 |
|  | After | 126199 (121.7%)* | 28644 (106.4%)* | 13459 (83.0%)* | 15185 (50.6%)* |
|  | Sum | 224633 | 55573 | 40068 | 15505 |
| 2017-2018 | Before | 107968 | 35570 | 16884 | 18686 |
|  | After | 52696 (48.8%)* | 9997 (28.1%)* | 6553 (57.6%)* | 3444 (38.8%)* |
|  | Sum | 160664 | 45567 | 23437 | 22130 |

Note: In 2019-2020, the time from the peak of the influenza positive rate to the Chinese first-level public health emergency response was 4 weeks. Accordingly, “the boundary point” was defined as 4 weeks added to the time when the influenza positive rate peaked in 2018-2019 and 2017-2018. The ILI and the positive influenza patients in the recent three years, before and after the boundary point were description in Table 2.
\*The percentages in brackets represent the ratio of the number after the boundary point to before the boundary point.

**Fig. 2.**
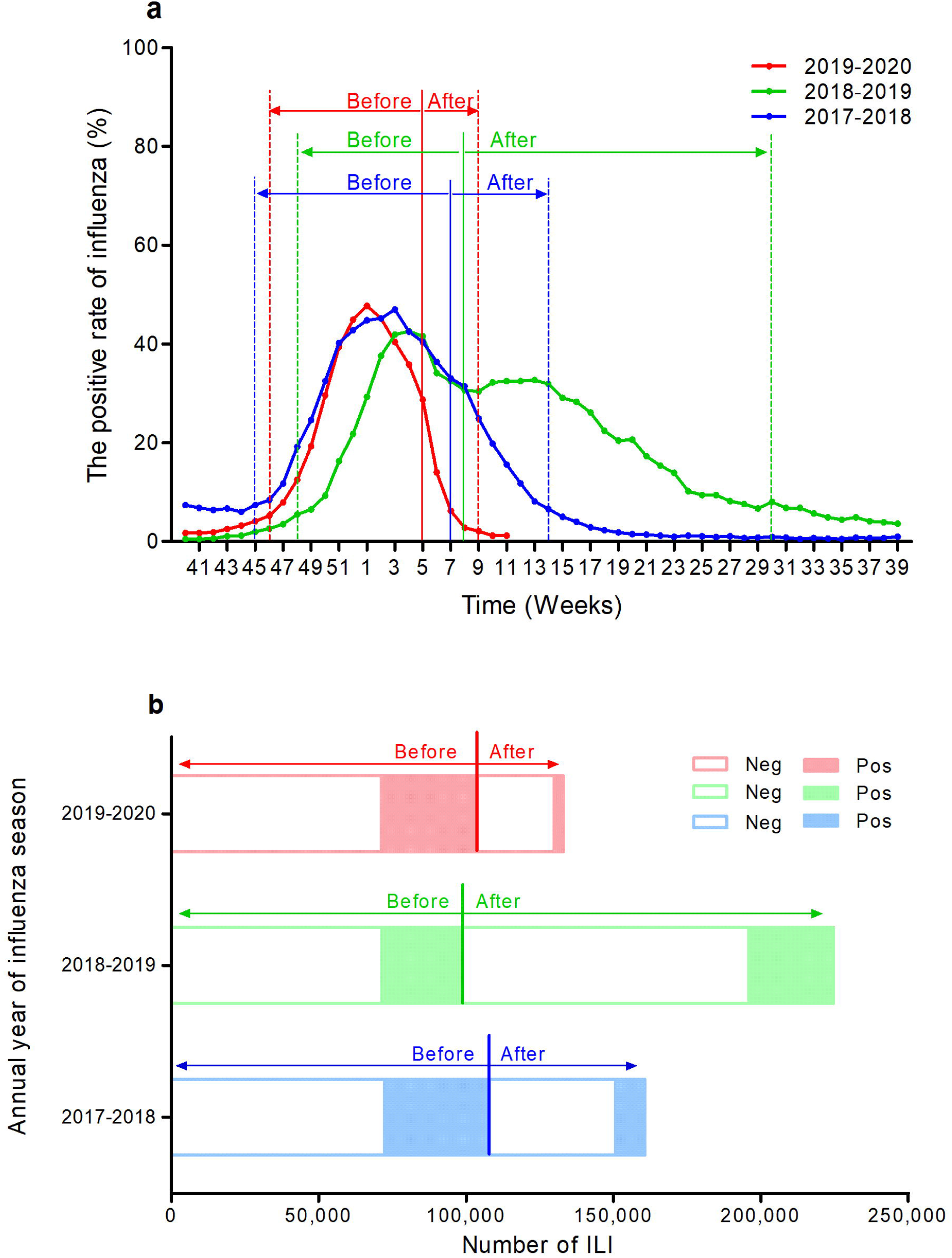
Comparing the trend of influenza epidemic in the recent three year. The time (weeks) from the peak of the influenza positive rate to the first-level response in 2019-2020 was calculated. Accordingly, the same number of weeks was added to the time when the influenza positive rate peaked in 2018-2019 and 2017-2018, from which the boundary point corresponding to the first-level response time in 2018-2019and 2017-2018 were calculated. (a) The weekly positive rates of influenza in the recent three years, before and after the boundary point were exhibited. The red, green, and blue vertical line represent the boundary point of 2019-2020, 2018-2019, and 2017-2018, respectively. (b) The ILI and the positive influenza patients in the recent three years, before and after the boundary point were exhibited. ILI, the influenza-like illness.

#### After initiation first-level response, with population mobility declining and the COVID-19 and influenza outbreaks contained

On 23 January, 2020, Chinese government issued a travel ban policy to lock down Wuhan and launched a first-level response to COVID-19. Then, the DNRP flow dropped rapidly, to 0.83 million on 13 February, which were only 30.2%, 13.6% and 9.5% during the first, second and third weeks after the Spring Festival in 2019, respectively. With DNRP flow dropped in entire China, the newly confirmed COVID-19 cases in mainland China except Hubei province showed a rapid decline after achieving the maximum number (890 cases) on 3 February. The newly confirmed cases were less than 50 cases per day since 21 February. Meanwhile, the prevalence of influenza also showed a significantly decline, with the positive rate dropping from 47.7% to 2.1% during the period from Jan 1 to March 1 of 2020 (Figure 1d).

## Discussion

COVID-19 broke out first in Wuhan, Hubei province in China and then quickly spread across the country. The first-level response was launched to control the spread of COVID-19. Coincidentally, it was just the Spring Festival travel rush in 2020. According to Chinese tradition, people will go back to home to celebrate new year coming. Huge population long distance mobility will extremely enhance virus rapid transmission. In this study, we focus on the measure-restriction of population flow, and analysis its relationship with COVID-19 and influenza.

The DNRP flow is used to reflect the population flow and indirectly evaluate the speed and effectiveness of the first-level response to the epidemic. During first-level response period, it was just the Spring Festival travel rush in 2020, with total 1.48 billion passengers sent by railways, highways, waterways and civil aviation, decreased 50.5% and 50.3% compared to the same period in 2019 and 2018 respectively. Since 23 January, the flow has decreased sharply (9.85 million on 23 January; 5.15 million on 24; 2.47 million on 25; fell to 0.83 million on 13 February). Chinese Spring Festival of 2020 was on 25 January, the DNRP flow in the first, second and third weeks after the Spring Festival were 26.6%, 13.0% and 8.7% of the same period in 2019, were 28.3%, 13.3% and 10.0%, respectively. According to the above results, it suggested that the first-level response had been implemented effectively.

Chinazzi M. et al. reported that restrictions on Wuhan city closure and international population movement had an impact on the spread of COVID-19 between China and other countries^9^. In fact, a series of prevention and control measures, such as controlling domestic population flow, have significantly inhibited the prevalence of COVID-19 in mainland China, along with the restrictions on the closure of Wuhan and the international population flow. After first-level response starting point (23 January), COVID-19 new cases in China had a slowly rise, then from 3 February, COVID-19 new cases outside Wuhan had been dropping. From 17 February, new cases of mainland China also began to keep falling. On 7 March, new cases were less than 100 in mainland China, and only 5 in Wuhan. The actual cumulative number of COVID-19 cases (excluding Wuhan) after 23 January was smaller than the epidemic curve of Wuhan without closure predicted by Chinazzi M. et al, and the difference was gradually significant with time^10^. Based on the modeling and analysis of 15 top research institutions in the world, Wuhan travel ban, as the largest isolation event in human history, combined with the first-level prevention and control measures, had reduced the number of Chinese patients with COVID-19 by more than 700,000 according the report of Tian et al^10^. Zhang YP’s study^11^ also showed that these measures may be conducive to control the epidemic. Therefore, we believe that prevention and control measures for the epidemic, including population movement restrictions, have played a significant role in the control of COVID-19.

Influenza, just like COVID-19, is also a respiratory infection disease. Data released by the influenza surveillance network laboratory in mainland China showed that A week before and after the first-level response, the influenza positive rate dropped from 40.4% to 35.8%, and at the last week of February, the influenza positive rate fell to 2.1%. And compared with 2018 and 2019, the influenza season was shortened in 2020. Although after reaching the peak in 2020, there has been a downward trend, the influenza epidemic trend in the United States, France and Italy were similar to previous years (Extended Data Fig. 1). As we know, the COVID-19 outbreak can be characterized as a “pandemic” as the virus spreads increasingly worldwide. China first-level response had strongly curbed the prevalence of COVID-19 in two months. It suggests the shorten influenza season in China might be related with the first-level response.

It is estimated that influenza could cause 3-5 million cases with severe respiratory infection-related illness and 0.29-0.65 million deaths worldwide annual year (Supplementary Table 2). Overall the mortality associated with influenza in China for adults aged 60 years or older were 38.5 per 100,000 person-seasonal, and for individuals younger under 60 years was 1.5 per 100,000 person-seasonal through 2010 to 2015^12^. According to the research of John Paget, overall rate of influenza-associated excess respiratory deaths for adults aged 65 years or older was 53.7 per 100,000 person-seasonal, while the mortality for individuals younger was 2.1 per 100,000 person-seasonal through 2002 to 2012^13^. The annual health economic burden associated with influenza should not be ignored^13-15^. The direct medical cost of out-patient cases of influenza is 156-595 RMB/person, and the indirect cost is 198-366 RMB/person. The economic burden of in-patient cases was about 10 times that of out-patient cases in China^16^. The estimated losses in 2015 will be about US $500 billion (US dollars) per year, and the economic losses caused by influenza will vary (0.3%-1.6%) due to differences in national economic conditions, accounting for about 0.6% of global income^13,14^. Therefore, the first-level responses not only prevent the spread of influenza, but also reduce the health economic burden caused by influenza that regularly occurs every year.

The first-level response also included the other measures, such as wearing face masks, washing hands and other good respiratory hygiene habits, avoid crowd gathering, symptoms or contact history of home isolation observation. As we know, hand hygiene is commended in most national pandemic plans^17^, and has been proven to prevent many infectious diseases. The face masks were common sight during the influenza epidemic period, but the role in preventing influenza virus transmission remains questionable^18^. Recently, Leung NHL et al’s research indicates that surgical face masks could prevent transmission of COVID-19 and influenza virus from symptomatic individuals^19^. Whether these measures play an important role in controlling COVID-19 and influenza need further research.

In summary, the first-level response had been observed effectively in controlling both COVID-19 and influenza epidemic. At present, COVID-19 has become the global pandemic situation^20^. Our study suggests if other countries adopt Chinese-style COVID-19 control policy, they would not only reduce the health and economic burden caused by COVID-19, but also reduce health and economic burden caused by other respiratory infectious diseases such as influenza. Therefore, when other countries issue measures on COVID-19, they should consider what they might also benefit from decreasing the influenza burden and other respiratory infectious disease by adopting more aggressive epidemic prevention and control strategies, just like China-style.

## Online Methods

### Collection of population flow data

To assess the first-level response for the prevention and control measures on the epidemic, we used the total passenger numbers sent in mainland China from 2018 to 2010 and the DNRP flow in 2020 during Spring Festival travel rush to reflect the population movement. The total number of passengers sent by railway, highway, waterway and civil aviation during the Spring Festival 2018 to 2020, and the daily number of passengers sent by railway during the Spring Festival 2020 obtained through the website of the Ministry of transport of China^21^.

### Surveillance of COVID - 19 data in China except Hubei province

In order to prevent and control COVID-19 in China, the NHC organized and updated the national diagnosis and treatment plan for COVID-19 timely. The new and cumulative cases of COVID-19 in China except Hubei province obtained from the NHC website^22^.

The definition of confirmed COVID-19 cases: 1. Nucleic acid of SARS-CoV-2 which detected by real-time fluorescence quantitative reverse transcription polymerase chain reaction (RT-PCR) was positive. 2. Virus gene sequence is highly homologous with SARS-CoV-2. 3. Specific IgM and IgG antibody of SARS-CoV-2 in serum were positive. Specific IgG antibody changed from negative to positive or 4 times higher or more in the recovery period than in the acute period. One of the above SARS-CoV-2 etiological or serological evidence was confirmed case^23^.

### Surveillance of influenza data

The NHC is responsible for influenza epidemic monitoring throughout the country, which refer to the national influenza surveillance plan (2017 version) ^24^. There are 554 National Sentinel Hospital and Influenza Surveillance Network Laboratories. Surveillance Subjects: ILI were with fever (body temperature≥38□), accompanied by cough or pharyngeal pain. Duration of surveillance: All National Sentinel Hospital for influenza case monitoring and Influenza Surveillance Network Laboratories conduct surveillance of ILI throughout the year. Reporting of ILI: National Influenza Surveillance Sentinel Hospitals and Influenza Surveillance Network Laboratories report their surveillance data to the CNIC before 24 o’clock on Monday. The ILI and influenza positive rate from October 2017 to February 2020 which required by this study were obtained from the weekly report of the CNIC^24^.

### Statistical analysis

All statistical tests were performed using SPSS 19.0 software. Categorical data was expressed as number (percentage), and Chi-square test was used for comparing difference between groups. Continuous data was expressed as the mean ± standard deviation 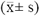, and t test or Mann-Whitney U test was used for inter-group comparison. A *P*<0·05 was considered to be statistically significant. The trend of population flow, COVID-19 and influenza epidemic was plotted using GraphPad Prism.

## Data Availability

The data used to support the findings of this study are available from the corresponding author upon request.

